# Founder effects arising from gathering dynamics systematically bias emerging pathogen surveillance

**DOI:** 10.1101/2022.11.15.22282366

**Authors:** Bradford P. Taylor, William P. Hanage

**Affiliations:** Center for Communicable Disease Dynamics, Department of Epidemiology, Harvard T.H. Chan School of Public Health, Boston, MA, USA

**Author notes:** **For correspondence:** (BPT).

## Abstract

Models of infectious disease transmission have shown the importance of heterogeneous contact networks for epidemiology; the most connected individuals are most likely to be infected early. Yet it is cumbersome to parameterize and incorporate such networks into simple models. We introduce an alternative model framework that explicitly includes attendance at and disease transmission within gatherings of different sizes, which disaggregates sequential epidemics moving from the most to least social subpopulations that underly the overall, single-peaked infection curve. This can systematically bias initial estimates of the growth rate for emerging variants and their severity, if vulnerable populations avoid large gatherings. Finally, we show that how often similarly social individuals preferentially interact (i.e., homophily, or assortative mixing) tunes the magnitude and duration of these biases. Together, we provide a simple framework for incorporating socialization and behavior in epidemic models, which can help contextualize surveillance of emerging infectious agents.

## Introduction

Directly transmitted infectious agents rely on the behavior of their hosts to start new infections and maintain themselves – specifically the contacts that their hosts make along which transmission may then occur. The contact networks that link individuals in a population are hence the routes along which communicable diseases spread, and considerable effort has been made to understand the nature of these networks and the implications for control of infectious agents (***Bansal et al., 2010***). The numbers of contacts individuals make can vary greatly, just as the nature of contacts can differ; some may be merely fleeting opportunities for a pathogen to initiate infection while others are sustained exposures to a large inoculum. Such heterogeneity in secondary contact rates appears to be common across epidemics, given over-dispersion of empirically observed secondary infection distributions, meaning that few individuals disproportionately contribute more towards ongoing transmission (***May and Anderson, 1987; Woolhouse et al., 1997; Lloyd-Smith et al., 2005; Stein, 2011***). While some of this may be due to infections with unusual properties, those infectious must still make contacts in order to transmit to them (***Quinn et al., 2000; Fraser et al., 2007; Matthews et al., 2005; Lawley et al., 2008; Gopinath et al., 2012; Edwards et al., 2021***).

Another feature that contact networks exhibit is assortativity: the tendency for contacts to be made between nodes with similar properties (***Newman, 2002***). In social networks this is termed homophily, and in the case of communicable disease the consequence is that most transmission events take place among groups of similar individuals, which can be explicitly incorporated with a contact matrix (e.g., for age-structured models) (***McPherson et al., 2001; Rohani et al., 2010***). Varying contact networks can be incorporated into dynamical models of transmission and have produced insights into subjects such as the optimal allocation of vaccines (those which prevent transmission are more effective when used in those with the most contacts) or the importance of core groups in maintaining sexually transmitted infections and as a focus of interventions (***Stigum et al., 1994; Endo et al., 2022***).

A special case of assortativity and homophily is when individuals share increased risks of exposure, and increased numbers of contacts – for example through gathering in groups of larger size. Larger gatherings both increase the chance that an infectious person attends, and the numbers to whom they may transmit (***Altizer et al., 2003; Godfrey et al., 2009; Chande et al., 2020***). The resulting variations in risk of exposure may result from voluntary social interactions (and contact rates tend to be higher in younger people), but can also include sexual networks in which the majority of at-risk contacts occur between a small proportion of the population, or crowded settings such as workplaces in which employees are unable to mitigate their own risks of exposure (e.g., meat-packing plants in the early stages of the covid pandemic are a well-known case of workplaces in which large outbreaks occurred) (***Yorke et al., 1978; Volz and Meyers, 2007; Taylor et al., 2020***). In the context of an emerging pathogen or variant such transmission heterogeneity and homophily mean that any estimates from local surveillance efforts may not reflect the epidemic potential in other communities. The social context of disease spread can produce biases in surveillance given differences in resources between settings.

Mathematical models may be used to project the expected consequences of outbreaks. For example, in the classic SIR model for a directly transmitted pathogen the basic reproduction number *R*_0_, determines the final size of the epidemic, but the model makes well known unrealistic assumptions about mixing patterns and must be extended to account for heterogeneity and homophily (***Kermack and McKendrick, 1927; Hébert-Dufresne et al., 2020***). Network models haven proven informative means to interrogate the effect of social structure on epidemic spread, yet by their nature they are demanding to parameterize (***Keeling and Eames, 2005; Keeling, 2005***). Nonetheless, capturing these heterogeneous contacts is necessary to contextualize the rapid growth of an epidemic as a result of the social context where the outbreak arises, rather than specific properties of the pathogen. This sort of “founder effect” is familiar in population genetics, but less so in epidemiology (***Mayr, 1942; Templeton, 2008***).

Here we present a simple extension of the classic SIR framework that specifically includes gathering sizes and homophily among individuals on the basis of the risk of infection. The resulting case curves are related to the gathering sizes and enable us to quantify how incorporating socialization alters our expectations for early spread of emerging viruses and variants and how this is reflected in seroprevalence and genomic surveillance.

## Results

### Transmission at gatherings: Group-SIR model

We propose modifying SIR models to account for varying contact rates and risk by explicitly modeling gathering dynamics using sampling processes. For any SIR-like model there are two key events, transmission and recovery, yielding infection dynamics:

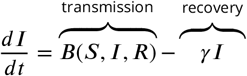

We focus on how gathering dynamics alter transmission, *B*(*S, I, R*) and ignore the effects of different recovery rates by normalizing time by the mean infectious period, 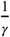. The rate gatherings occur, *r*_gather_, along with the size of the gatherings determine the social component of transmission that is controllable by non-pharmaceutical interventions. At a gathering, transmission can occur between each pair of the *n* attendees who were randomly sampled from the overall population. Figure 1a illustrates this sampling process for the classic SIR model where n=2 and Figure 1b illustrates this for the generalized “group-SIR” model. The expected number of infections at a gathering determines the overall transmission rate:

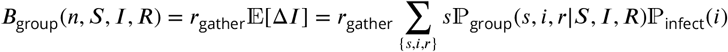

with probabilities

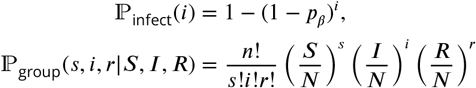

where lower-case *s, i*, and *r* denote the integer number of sampled susceptible, infectious, and recovered individuals attending the gathering and uppercase *S, I, R* denote the population-level densities such that *S* + *I* + *R* = *N*. To average, we sum over all possible combinations of *n* SIR attendees sampled from the population densities according to a multinomial distribution, ℙ_group_. A susceptible is infected at the gathering with probability, ℙ_infect_ whenever any attending infectious individual transmits according to a pairwise probability of infection, *p*_*β*_. The dynamics simplify to a classic SIR model when gatherings are restricted to pairs, n=2, whereas the transmission term becomes increasingly nonlinear for larger gatherings (see Appendix for details).

**Figure 1.**
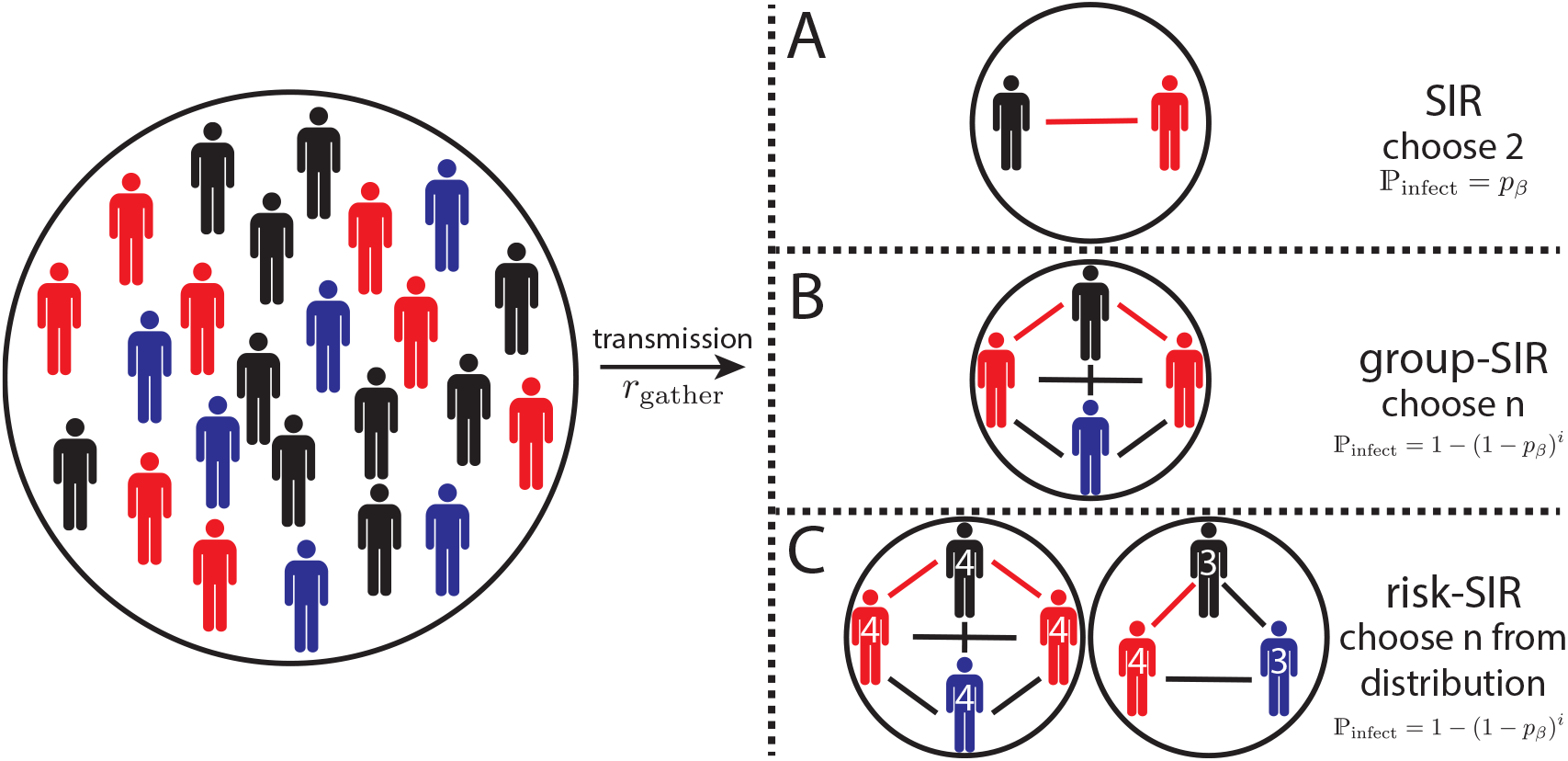
Schematic of sampling processes leading to different epidemic models described. A) The classic SIR model arises from sampling 2 individuals from a population of susceptible (black), infectious (red), and recovered (blue) individuals. B) Sampling *n* individuals yields a generalization of the SIR model, the group-SIR. Infections occur independently between each S-I pair present in a congregation. C) Denoting individuals by the maximum congregation size they are willing to attending yields the risk-SIR model. Societies can be defined by a distribution of congregation sizes which determine the epidemiological dynamics (***Boyer et al., 2022***).

Increasing gathering sizes leads to sharper epidemics for otherwise fixed parameter values (Figure 2a). This is expected since increasing group sizes implicitly increases the total number of contacts relative to the recovery rate, i.e., increasing the basic reproduction number, *R*_0_. Rescaling the transmission rate by the total number of pairwise interactions at a gathering, 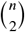, collapses the group-SIR dynamics onto each other (Figure 2a inset). There are fewer cases at the epidemic peak for larger gatherings as a result of multiple infectious individuals infecting the same susceptible, effectively reducing the number of pairwise interactions. In short, congregating in larger groups increases the rate of epidemic spread, but has minor impact on the dynamics when correcting for the number of interactions except by reducing the cumulative number of infections, in line with prior observations (***Volz et al., 2011***).

**Figure 2.**
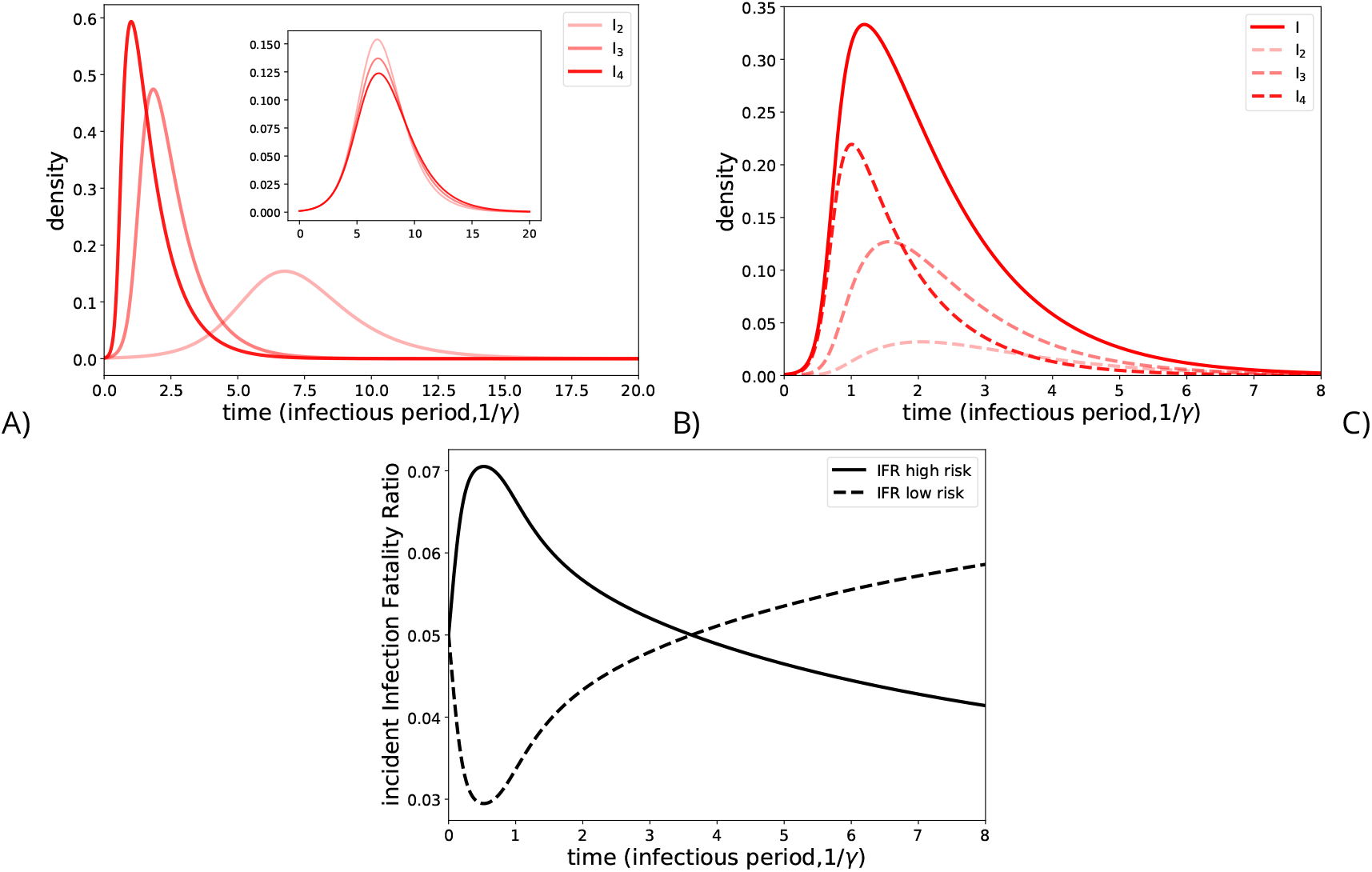
Generalized SIR dynamics. A) Group-SIR dynamics, where transmission occurs within groups with sizes here denoted by the subscript. B) Risk-SIR dynamics, where transmission occurs within groups according to a distribution here set with sizes n=2,3, and 4 at equal frequency. Subpopulations are denoted by their risk level shown, which specifies the maximum gathering size they are willing to attend. C) The sequential peaks of infection cause the overall infection fatality ratio (IFR) to be dynamic when it varies between subpopulations. Here we compare IFR dynamics when the IFR is positively correlated with risk level (IFR high risk) and when the IFR is negatively correlated with risk level (IFR low risk). Note, we assume that mortality dynamics are the same as recovery dynamics to demonstrate this qualitative point.

### Gathering size dependent attendance: Risk-SIR model

During infectious disease outbreaks individuals may vary in how much they alter their behavior in response to perceived risk (***Perra et al., 2011; Eksin et al., 2017; Arthur et al., 2021; Harris et al., 2023***). To capture this we can further generalize the group-SIR dynamics where individuals choose among differently sized, concurrent gatherings and avoid those larger than their “risk level”:

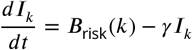

where risk levels are denoted by subscripts. Transmission follows from repeatedly sampling from the population for a given distribution of differently sized gatherings, ℙ_*n*_;

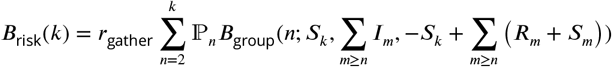

Each transmission term captures new infections within a particular focal subpopulation of individuals with the same risk level, *k*, such that only the respective susceptibles, *S*_*k*_, contribute. These focal susceptibles can be infected when attending any gathering no larger than their risk level, *n* ≤ *k*, by any attending infectious individual with risk levels at least as large as the respective gathering size, *m* ≥ *n*. In contrast, infections among susceptibles from other subpopulations do not contribute to new focal infections and thus they function similarly to the recovered population. Overall, the transmission term is a weighted average over all gathering sizes according to a distribution, *P*_*n*_, set by society (e.g., by lockdowns) (***Boyer et al., 2022***). The risk-SIR model includes transmission heterogeneity as subpopulations with higher risk level attend a larger fraction of gatherings, homophily as subpopulations with higher risk levels preferentially interact at larger gatherings, and super-spreading as multiple infections can occur at large gatherings.

The risk-SIR yields single-peaked epidemics like the classical SIR, but with underlying sequential peaks among decreasing risk level subpopulations (Figure 2b). Differences in growth rates cause these different peaks, namely that susceptibles in higher risk subpopulations attend larger gatherings on top of attending the same gatherings at the same rate as those in lower risk subpopulations. Similarly, infections in higher risk level subpopulations lead to more secondary infections. The basic reproduction number of an infectious individual with a specified risk level, *k*, follows a recurrence relation:

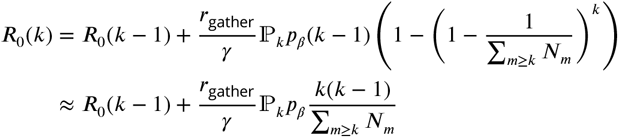

such that a disproportionate number of secondary infections occur at larger gatherings as enumerated by *k* (see Appendix for derivation). The largest fraction of the secondary infections occur within the same risk subpopulation or in larger risk subpopulations as these susceptibles can attend all the same gathering as those infectious, whereas susceptibles in smaller risk subpopulation do not attend the larger gatherings. Together, these dynamics cause the initial growth of the epidemic to be driven by the largest risk subpopulations, in line with prior work (***May and Anderson, 1987***).

### Impact on Surveillance

The sequential epidemic peaks within risk subpopulations have multiple impacts for disease surveillance. First, the cases initially accumulate within subpopulations at most risk, highlighting the importance of caution when interpreting and designing serosurveys to account for differential risk of exposure. Any underlying surveillance metrics that vary between subpopulations will now be dynamic. For example, we use infection fatality rates (IFR) to predict the threat of an emerging outbreak. If virulence depends on comorbidities that vary in frequency between each subpopulation then the IFR will change over the successive peaks of the underlying outbreaks in different sub-populations. Figure 2c illustrates IFR dynamics for two general cases. First, when individuals with comorbidities choose to avoid larger gatherings, i.e., subpopulation IFR negatively correlates with risk level (IFR low risk), then the IFR increases over time because at risk populations are infected relatively more frequently at later times. This would also reduce case ascertainment at the outset of an epidemic, potentially biasing for lower estimates of *R*_0_. Conversely, if individuals with comorbidities attend larger gatherings, i.e., subpopulation IFR positively correlates with risk level, then the IFR decreases over time (IFR high risk). This latter example is particular relevant for essential workers during lockdowns and communicable childhood diseases within school settings. IFR dynamics emphasize the danger in reporting aggregate statistics without accounting for disproportionate disease spread among subpopulations, such as when tracking emerging variants.

An epidemic grows at a rate determined by the socialization of all infectious individuals, i.e., it’s risk distribution. Meanwhile, a variant emerges within a subset of these infectious individuals. Thus, even a neutral variant with no increased transmissibility relative to the background it emerges in is expected to increase in frequency if it is transmitted by individuals more social than average across the epidemic. Figure 3a shows the risk distribution of an epidemic and a neutral variant emerging within the most social subpopulation at a given time. The resulting variant frequency dynamics, shown in Figure 3b, mimic logistic growth–approaching an equilibrium frequency prior to sweeping. This dynamic emphasizes the role that social context plays in determining the observed growth rate of a variant. For example, a variant of concern with increased transmissibility relative to wild type will ultimately grow in frequency, but if it emerges within a less social subpopulation then the observed increase in frequency will be delayed (figure 3c) increasing its chance to stochastically become extinct. The same plot with a logarithmic y-axis shows a clear decreases in frequency of the more transmissible variants that emerge in less social subpopulations, which increases the chance for stochastic extinction (See Appendix figure 1).

**Figure 3.**
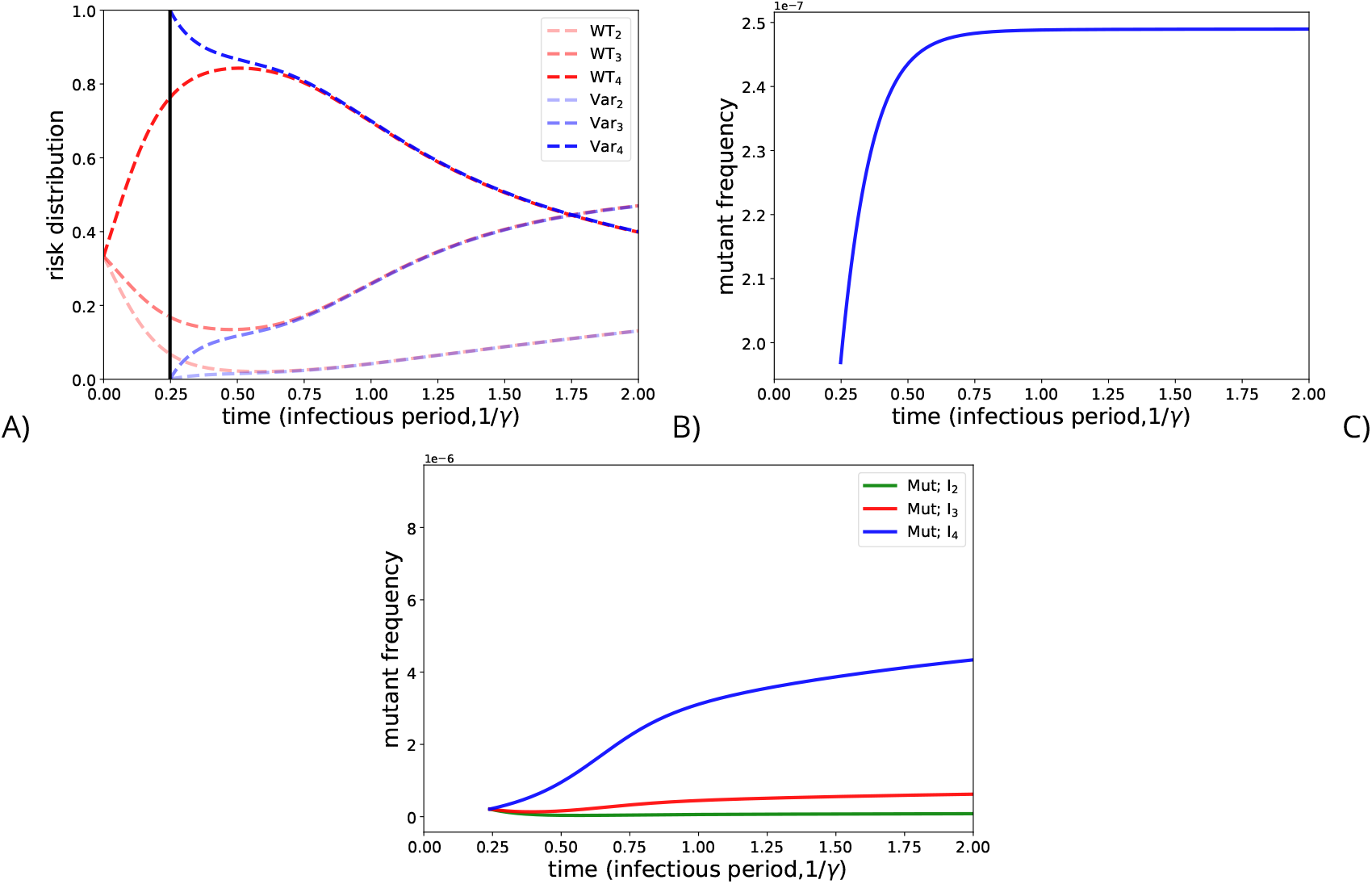
Variant frequency dynamics depend on which risk subpopulation it emerges within. A) Risk distribution of an epidemic (red) and a neutral variant (blue) that emerges within the highest risk subpopulation. B) The associated frequency dynamics of the emerging neutral variant. C) Frequency dynamics of a variant with 50% greater transmissibility relative to the background epidemic emerging in different risk subpopulations

Since both increased transmissibility and increased contact opportunities contribute to the apparent fitness of a variant, identifying fast growing lineages during genomic surveillance imposes a selection bias towards identifying lineages preferentially spreading among more social individuals. To see this, we simulated the individual transmission and recovery events of a risk-SIR model using the Gillespie algorithm and tracked all possible lineages (***Gillespie, 1977***). Figure 4a shows the frequency dynamics of the fastest growing lineage (having reached a given size at a given time - dotted vertical line). This lineage is neutral but appears to be growing more rapidly than its peers, and continues to increase in frequency after identification. This occurs because the risk distribution of the variant at the point of sampling, shown in Figure 4a (inset), contains relatively more social individuals relative to the rest of the epidemic and, in turn, an expected increase in frequency relative to the epidemic. Note, here and throughout we follow common usage by epidemiologists to refer to pathogen clades as lineages, i.e., all ancestors of a focal case.

**Figure 4.**
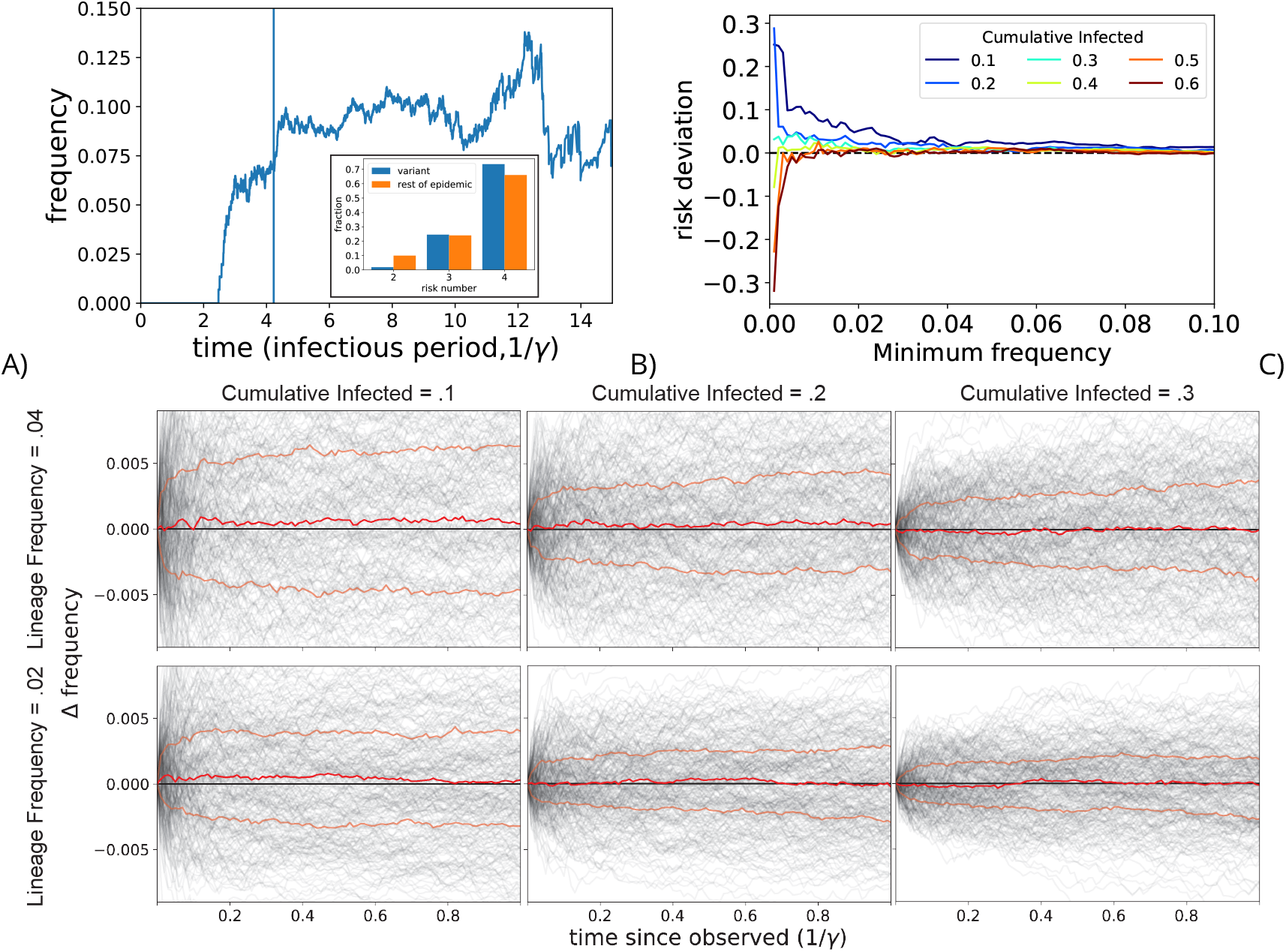
Selection bias of fast growing clades a) An example of a fast growing clade that continues to grow beyond the point it was identified (blue vertical line) and its risk distribution among infectious individuals as compared to all others infectious at the moment of censusing (inset). b) Median difference between the median of the fastest growing risk distribution and the risk distribution of the rest of the epidemic for given censusing frequency within a 100% bandwidth for epidemics with subpopulations [5000, 0, 5000, 0, 5000] for risk numbers 2-6. c) Relative frequency dynamics of the fastest growing lineages following censusing across different censusing parameters for epidemics with subpopulations [5000, 0, 5000, 0, 5000] for risk numbers 2-6. Red lines denote median and interquartile ranges across 250 simulations shown in transparent black lines.

Whether identifying fastest growing lineage biases for those with spreading among more risky subpopulations depends on censusing parameters. This dependence arises because selecting fast-growing lineages biases the risk distributions by two opposing processes. On one hand fast-growing lineages are more likely to spread among the riskiest subpopulations because they attend more and larger gatherings and subsequently more frequent secondary infections. On the other hand, the fastest growing lineages preferentially include recent infections, which bias towards less risky populations given the sequential spread of the epidemic from more risky to less risky subpopulations over time. Figure 4b shows the the median risk distribution bias for different censusing parameters across 250 repeated simulations of an epidemic spreading within a society with high variance in gathering rates (see caption). The x-axis specifies the minimum frequency within a 100% bandwidth that the fastest growing lineage will be tracked, e.g., 0.01 refers to the fastest growing lineage among those between .01 and .02 frequency. Fast growing lineages tend to have biased risk distribution particularly early on during epidemics, in line with greater stochastic deviations of smaller lineages for a given frequency. Figure 4c shows whether the identified lineages tend to grow over time due to the bias for different censusing parameters. We see that the long-term effect of the bias is larger for moderate frequencies (.04 vs .01), due to the fact that larger lineages for a given frequency take longer to deterministically relax their risk distribution to that of the entire epidemic. Note, given the impact of this bias is larger for moderately sized fast-growing lineages means we are susceptible to identifying these lineages during surveillance. For example, the CDC tracks lineages > .01 frequency. Our above results suggest these lineages are susceptible to being misidentified as having increased transmissibility and that their continued transient increase in frequency following censusing would incorrectly solidify these concerns. By understanding how social structure modulates these biases, we can better establish from which communities we expect to identify these spurious variants of concern.

### Society modulated surveillance bias

Who interacts with whom in a society affects the magnitude of the above identified surveillance biases. For example, the amount of homophily, i.e., assortative mixing, within a community sets the time scale over which such lineages will continue to spuriously increase in frequency. Intuitively, homophily increases the probability a skewed risk distribution remains skewed as onward infections primarily occur within riskier subpopulation. As the risk-SIR model has a fixed social structure and homophily, we can better understand the relationship between homophily and skewed risk distribution retention by analyzing a simplified SIR type model that disentangles transmission heterogeneity and homophily:

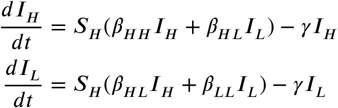

where ***H*** denotes highly social individuals and ***L*** denotes less social individuals. This is a minimal model including transmission heterogeneity when the subpopulations differ in their basic reproduction number, i.e., 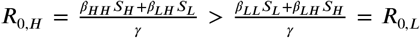, while smaller values of cross-transmission, *β*_*HL*_, correspond to societies with greater homophily. For this model, the risk distribution simplifies to the relative fraction of the more social subpopulation 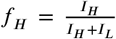. We can solve for the equilibrium risk distribution, 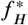, early on in an epidemic during exponential growth by fixing the susceptibles (*S*_*H*_ and *S*_*L*_) to constants and solving the respective dynamics, 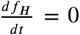.The equilibrium risk distribution depends on the structure of society, with increasing homophily reflected as a higher proportion of highly social individuals (see Appendix figure 2).

Randomness in individual transmission and recovery events (i.e., demographic stochasticity) causes the risk distribution to drift, even when at equilibrium. We can solve for the dynamics of the probability distribution over risk distributions as induced by demographic stochasticity via a so-called “master equation” parametrized by the average rates of the individual events (***van Kampen, 2007***). Figure 5a shows probability distributions after 500 population changes (transmission or recovery) for epidemics varying in homophily and initiated with 20 infectious individuals distributed according to their respective equilibrium risk distribution. Note, we vary homophily alone and keep the same levels of transmission heterogeneity across models by fixing the subpopulation basic reproduction numbers, ***R***_0,*H*_ and ***R***_0,*L*_. Demographic stochasticity induces a spread that eventually dissipates over time (see Appeneix figure 3). This means emerging variants are likely to deviate further from the equilibrium growth rate than the rest of the epidemic. This motivates quantifying how often a variant will grow faster versus slower than the equilibrium growth rate, 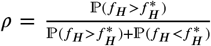. Figure 5b compares the relative probability of increased sociality, *ρ*, dynamics after initiating infectious populations at their respective equilibrium value,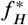, for different amounts of homophily. Emerging variants in societies with increased homophily have larger bias towards increased growth rates relative to the overall epidemic. Figure 5c shows the bias in risk distribution for epidemics initiating either in the more social (***I***_*H*_, dashed lines) or the less social (***I***_*L*_, solid lines) subpopulations. The subpopulation in which a variant emerges sets an initial risk distribution bias and societies with increased homophily retain this bias longer. However, the risk distribution bias ultimately approaches the same biased quasi-equilibrium value as seen above when started at the equilibrium.

**Figure 5.**
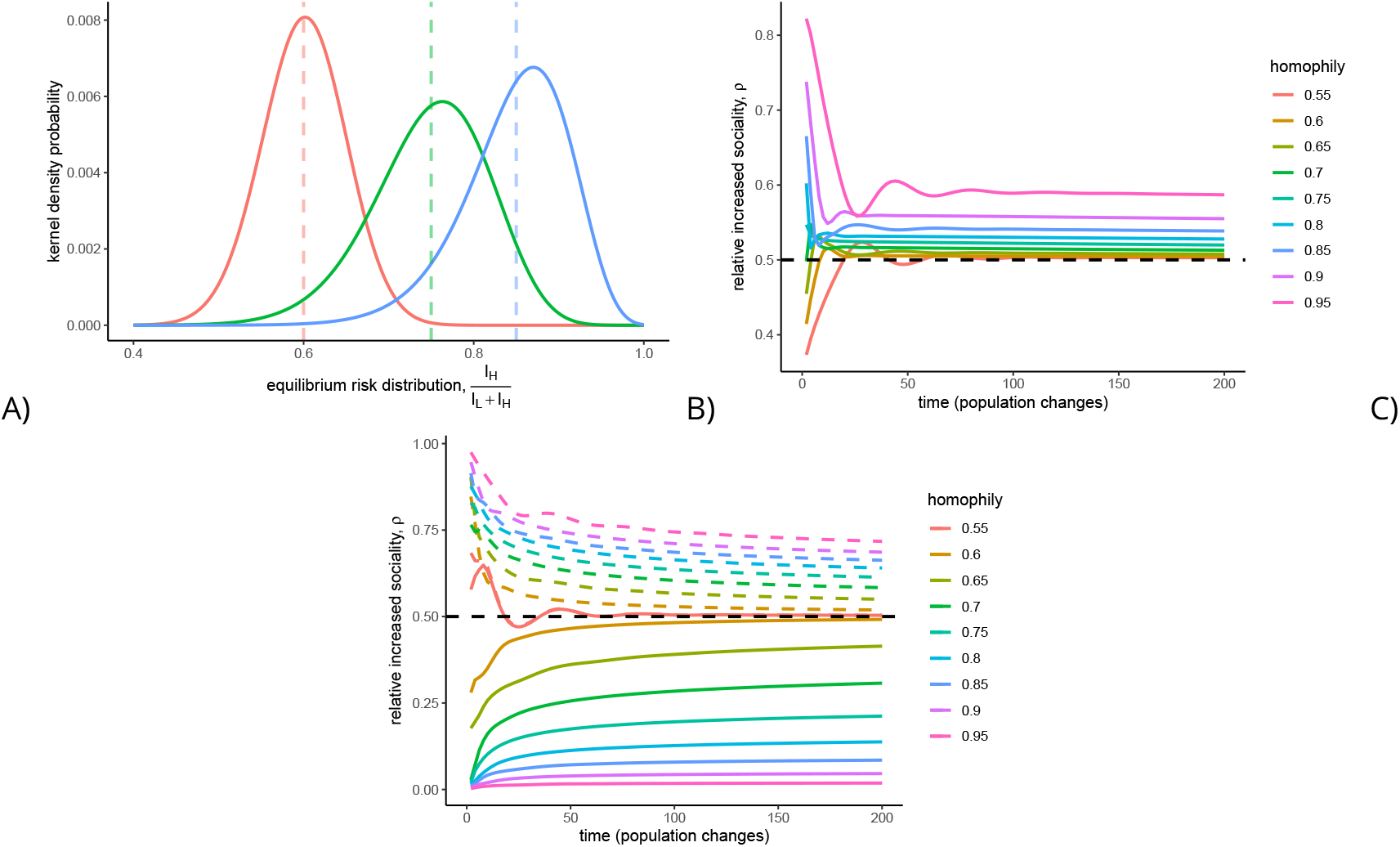
Homophily biases risk distributions a) Smoothed probability distributions of the risk distribution of epidemics initiated with 20 individuals at the respective equilibrium distribution after 500 population changes (total recovery or transmission events). Vertical dashed lines specify respective equilibrium risk distributions. B) Relative probability of a more social risk distribution 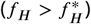 than a less social risk distribution 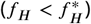 for epidemics initiated with 20 infectious individuals distributed according to the equilibrium risk distribution 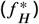 denoted as homophily. C) Relative probability of a more social risk distribution 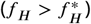 than a less social risk distribution 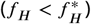 for epidemics initiated with 1 infectious individuals either in the more social subpopulation (dashed lines) or the less social subpopulation (solid lines).

Preferential stochastic extinction of slower growing epidemics exacerbates the growth bias induced by homophily. An epidemic goes stochastically extinct when its last remaining infectious individual recovers prior to onwards transmission. Intuitively, epidemics are more likely to go stochastically extinct when the last remaining infectious individual is among a less social subpopulation. This further increases the growth bias beyond that induced by homophily alone (see Appendix Figure 4). Hence, we expect emerging variants to increase in frequency by preferentially spreading among more social individuals, particularly when they emerge in societies with high levels of homophily.

## Discussion

The dynamics of communicable diseases are intimately linked to the contacts that are made between their hosts (***Buckee et al., 2021***). These are the opportunities to transmit and initiate new infections, and must be accounted for if we wish to interpret recent data during the course of an outbreak or forecast its course. Here we have presented a simple and flexible means to incorporate network structure implicitly into the classic SIR-framework and relate it to varying risks of exposure. By allowing interactions within groups of size > 2, representing varying risk-groups as a result of choice or necessity, the dynamics can no longer be described simply as the product of the proportions of the population that are susceptible or infectious, allowing us to examine the dynamics in separate risk groups and the impact on individual chains of transmission within them.

Prior studies on how contact heterogeneity impacts disease spread has been a major contrib-utor to the epidemiology of HIV-1 among at-risk populations, and more recently Mpox transmission along dense contact networks among men who have sex with men (***May and Anderson, 1987; Endo et al., 2022***). This includes sequential spread to populations with low contact rates from ‘core groups’ characterized by higher numbers of contacts (***Yorke et al., 1978; Colgate et al., 1989***). Our work explicitly includes gatherings as a mechanism for the separation of and homophily between high and low contact subpopulations. This context is more relevant for pathogens with shorter latency time than HIV including respiratory viruses like SARS-CoV-2 in which super-spreading (i.e., overdispersed transmission) plays a large role in the observed dynamics – especially at the out-set as the pathogen is introduced and becomes established. By emphasizing the importance of gatherings as the locus of spread, our model is aligned with interventions that utilize retrospective contact tracing to enable epidemiologists to monitor key locations where transmission occurs and link these data to population-level predictions (***Boyer et al., 2022***).

In an unmitigated outbreak, the only factor reducing the rate of new infections is convalescent immunity, and it has been recognized for some time that variable contact rates can have a large impact on the point at which this begins to take effect (***Hébert-Dufresne et al., 2020***). This is important for vaccination strategies aimed at those most likely to become infected and transmit, as it can markedly reduce the ‘herd immunity threshold’ necessary to achieve control and prevent large outbreaks (***Woolhouse et al., 1997***). Our model readily captures this and shows moreover that a single outbreak is itself made up of multiple ones with different dynamics in distinct but overlapping networks with distinct but overlapping epidemic curves. Those with the most contacts are rapidly infected, leading to the counterintuitive result that those who make the fewest contacts and/or have the fewest risks of exposure are more likely to be infected at times when the force of infection is lower, particularly late in the epidemic. This finding is important when interpreting the relationship between crude case counts and severe outcomes, where the latter vary markedly among individuals. It is illustrated by the use of hospitalizations and deaths as a lagging indicator of cases early in the COVID-19 pandemic, and likely contributes to the much smaller variance in numbers of severe outcomes over time as compared with estimates of case counts or wastewater data (***Khan et al., 2023***). Intuitively, those who are most vulnerable are least likely to be infected early on, but they and their networks will remain capable of supporting a sustained outbreak over a longer period of the epidemic.

This can also produce substantial bias in the apparent value of key parameters that are important for forecasting and estimating the impacts of an outbreak. If there is any correlation between the risk groups and the probability of severe illness, quantities such as the infection fatality rate or infection hospitalization rate will be biased. And this bias is in addition to those already known to arise through under-ascertainment of mild infections (***Accorsi et al., 2021***).

The model also suggests caution when interpreting and estimating the fitness of emerging lineages such as putative ‘variants of concern’. The founder effect in population genetics refers to a spurious appearance of fitness as a result of stochastic factors, as when a small number of migrating individuals colonizes a deserted island (***Wright, 1942***). Here, it indicates that early on in an epidemic the initial growth rate of any lineage that becomes common enough to be noticed is expected to be biased upward in relation to its true fitness, simply because it is spreading by definition in the networks which make the most contacts. Conversely, as the overall epidemic is declining those lineages that are persisting are infecting a higher proportion of fewer contacts, and so past the peak increases in proportion of a sample becomes a more reliable indicator of fitness. Because we have developed an extension of the SIR model, we have not explicitly considered immune evasion. However it can readily be seen that the host population in which a pathogen with immune evasion is most likely to experience both high fitness and start spreading is that which makes the most contacts and within which seroprevalence is highest (***Bushman et al., 2021***). Although this may be complicated by the timescale of waning immunity this is beyond the scope of the present work.

The SIR model has been a valuable source of intuition for generations of epidemiologists but is known to be limited. Here we have shown the value of a small change to incorporate larger group sizes than pairwise interactions. The results help interpret the expected course of epidemics where socialization and awareness of the epidemic drive transmission, such as during the recent Mpox outbreak (***Endo et al., 2022***). Furthermore, our model highlights how varying exposure leads to under-appreciated sources of bias when tracking putative variants of concern. Overall, the simplicity of the model may allow it to be readily applied to more complicated scenarios in which risk perception varies over time and the relative compositions of risk groups change accordingly.

## Data Availability

All data produced are available online at https://github.com/bradfordptaylor/risk_sir

## Code availability

All code used to generate the figures is available at: https://github.com/bradfordptaylor/risk_sir.

## Acknowledgements

This project has been funded (in part) by contract 200-2016-91779 with the Centers for Disease Control and Prevention and by contract U01 CA261277 through SeroNet. Disclaimer: The findings, conclusions, and views expressed are those of the author(s) and do not necessarily represent the official position of the Centers for Disease Control and Prevention (CDC).

## Appendix

### *n* = 2 and *n* = 3 group-SIR transmission terms

Here, we expand the group-SIR transmission term to connect it to the classic SIR model and to show how transmission grows nonlinearly with increasing group size. The sampling process yields transmission rates proportional to the expected number of infections at gathering with a given size *n*:

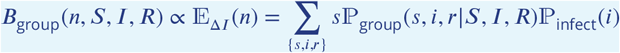

with probabilities

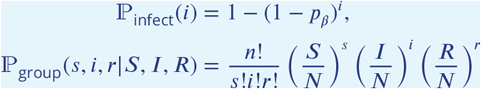

Transmission only occurs at gatherings when at least one infected and at least one susceptible attend. Hence, only one combination of attendees contributes in the *n* = 2 case:

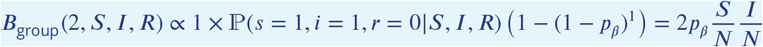

which is proportional to the bilinear classic SIR model. Note, the emphasis on proportionality, as this from differs from standard presentation of the SIR transmission by a constant factor of 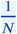 which can be accounted for by the rate of gathering *r*_gather_. The *n* = 3 case includes transmission with more combinations of *s,i*, and *r* attendees:

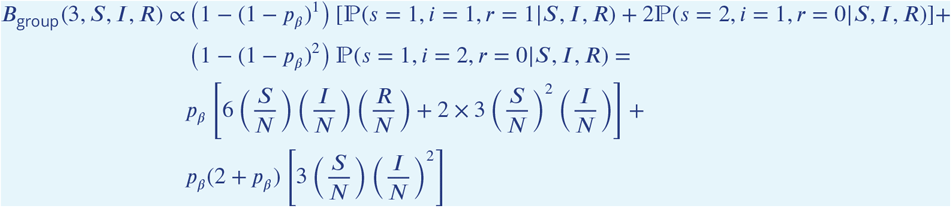

which deviates from a classical SIR model due to higher order nonlinearity.

### risk-SIR R_0_ derivation

The basic reproduction number, *R*_0_, is defined as the average number of secondary infections transmitted by an index case in an otherwise uninfected population. This can be calculated from the dynamics of gathering and the expected number of infections given the combinatorics of attendance. Because individuals with different risk levels attend different gatherings, we calculate the basic reproduction number for each risk subpopulation. Three terms contribute to *R*_0_(*k*): (1) the total number of gatherings at given sizes that an individual with risk level *k* can attend while infectious, (2) the probability that the individual attends each gathering, and (3) the average number of infections at each gathering given the infectious individual attends. The average number of gatherings for a given size *n* ≤ *k* that occur during the average infectious period is:

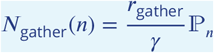

where *r*_gather_ is the rate all gatherings occur, *γ* is the recovery rate, and ℙ_*n*_ is the probability a gathering is size *n*. The probability an individual with risk level ≥ *n* attends a gathering of size *n* is:

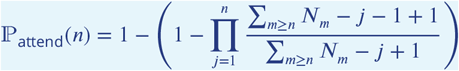

which is the complement of the probability an infectious individual not being sampled as any of the *n* attendees from the total individuals with sufficiently high risk levels, *N*_*m*_. The expected number of infections at a gathering of size *n* when the infectious individual attends is simply:

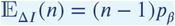

where *p*_*beta*_ is the pairwise infection probability. We combine these terms, sum over all possible gathering sizes that the infectious individual with risk level *k* is willing to attend, and simplify ℙ_attend_ in the limit of infinite population sizes, *N* → ∞, to get the final result:

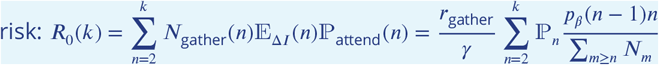

We show this result as a recurrence relation in the text to highlight how it scales with *k*.

**Appendix 1— figure 1.**
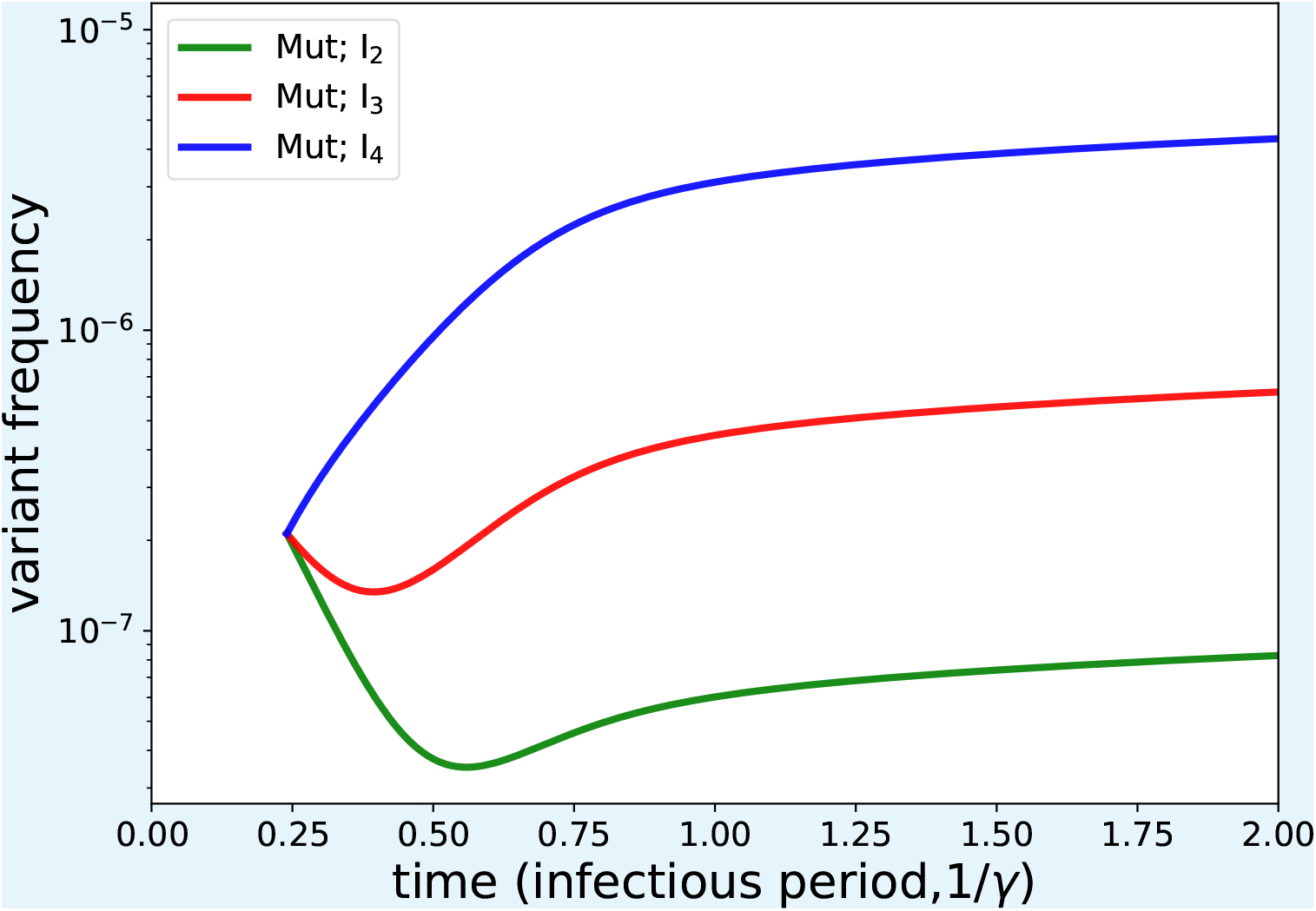
Dynamics of a variant with increased transmissibility relative to wildtype emerging in different risk subpopulations.

**Appendix 1— figure 2.**
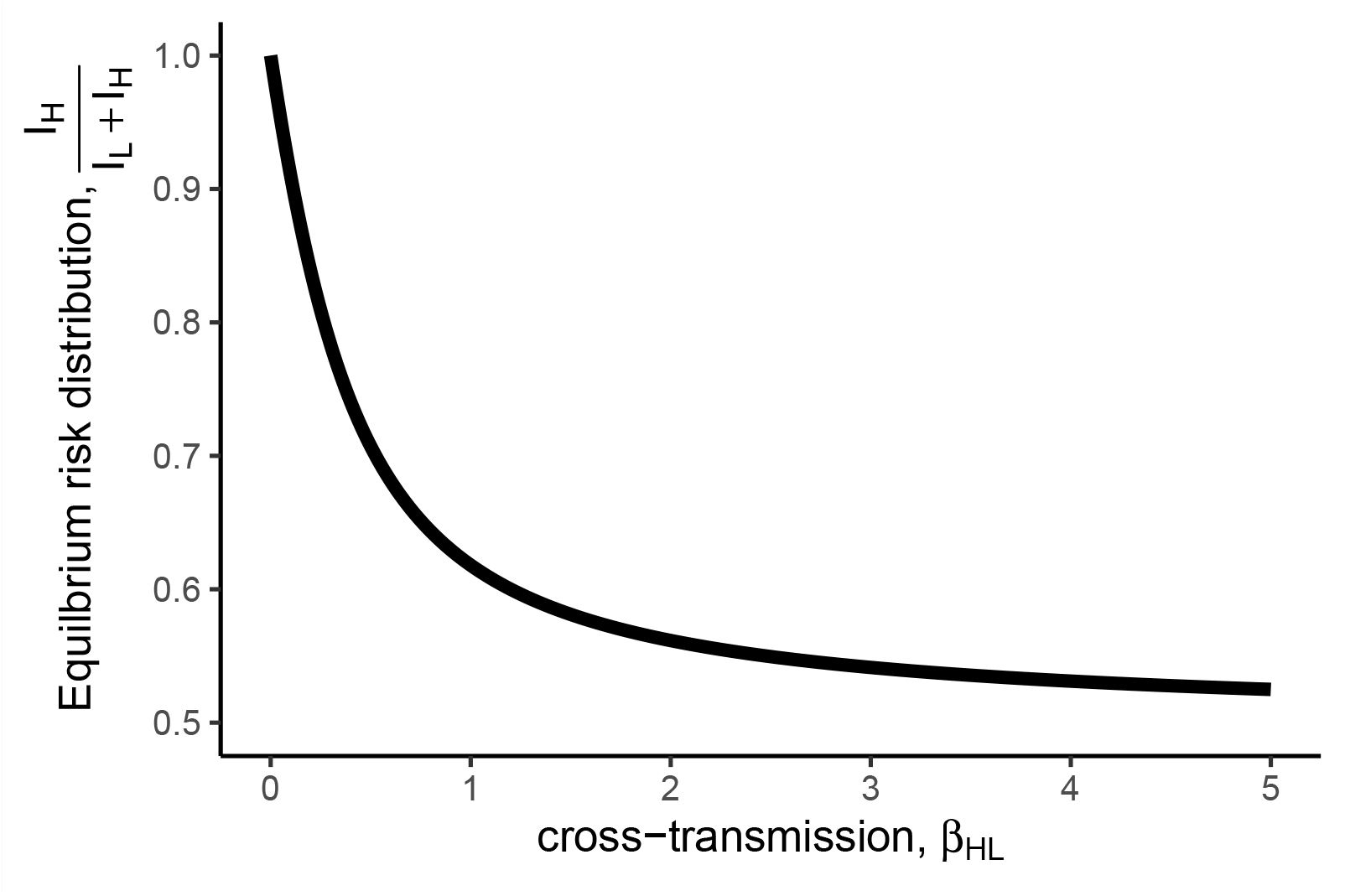
Equilibrium risk distribution for exponentially growing epidemics in societies with transmission heterogeneity as the amount of cross transmission between two subpopulation varies.

**Appendix 1— figure 3.**
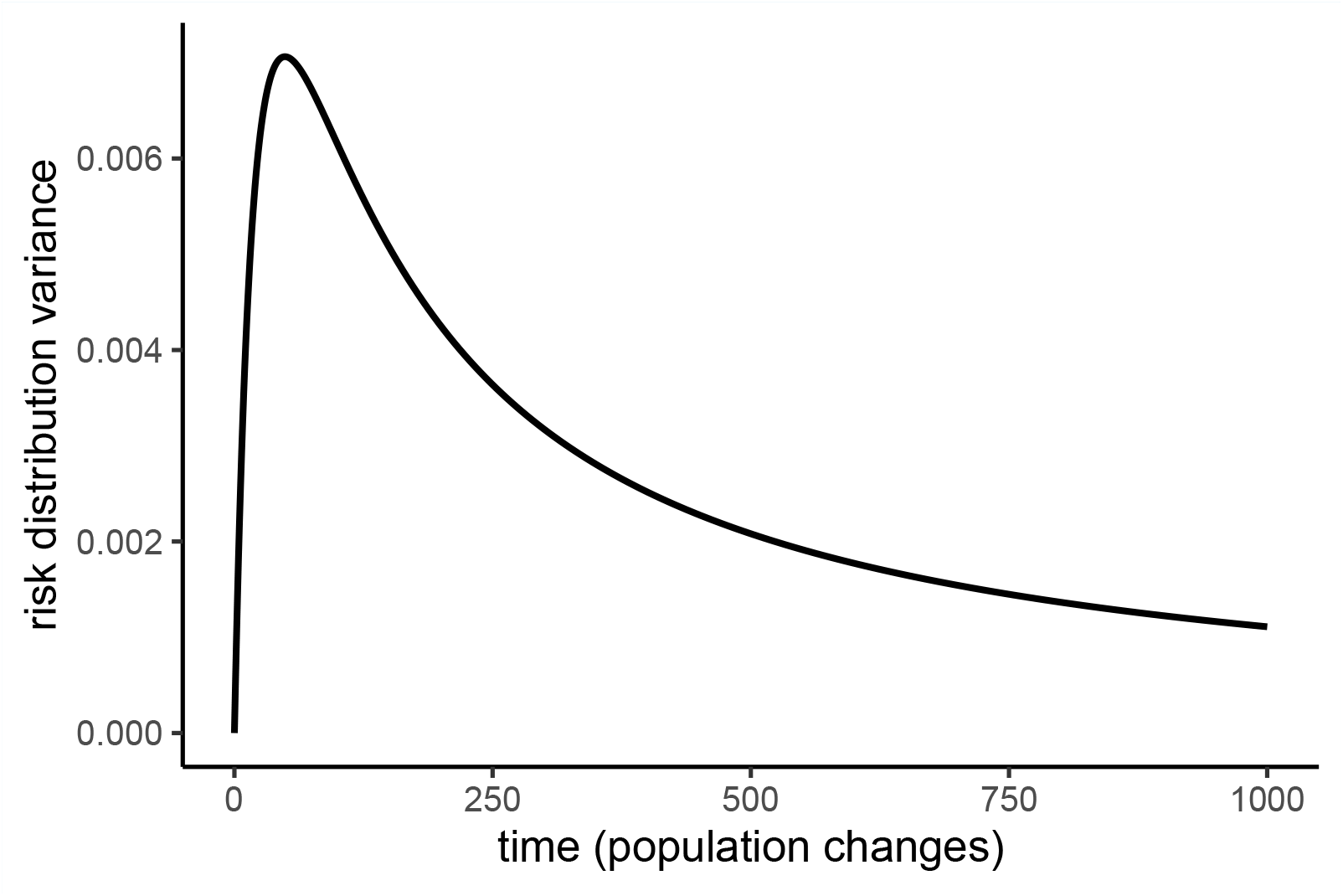
Dynamics of the variance of the probability density function over risk distributions for a growing epidemic with two subpopulations. It initially increases because we initialize the population at a specific risk distribution (delta distribution) and it eventually decreases as the risk distribution drifts less for larger populations.

**Appendix 1— figure 4.**
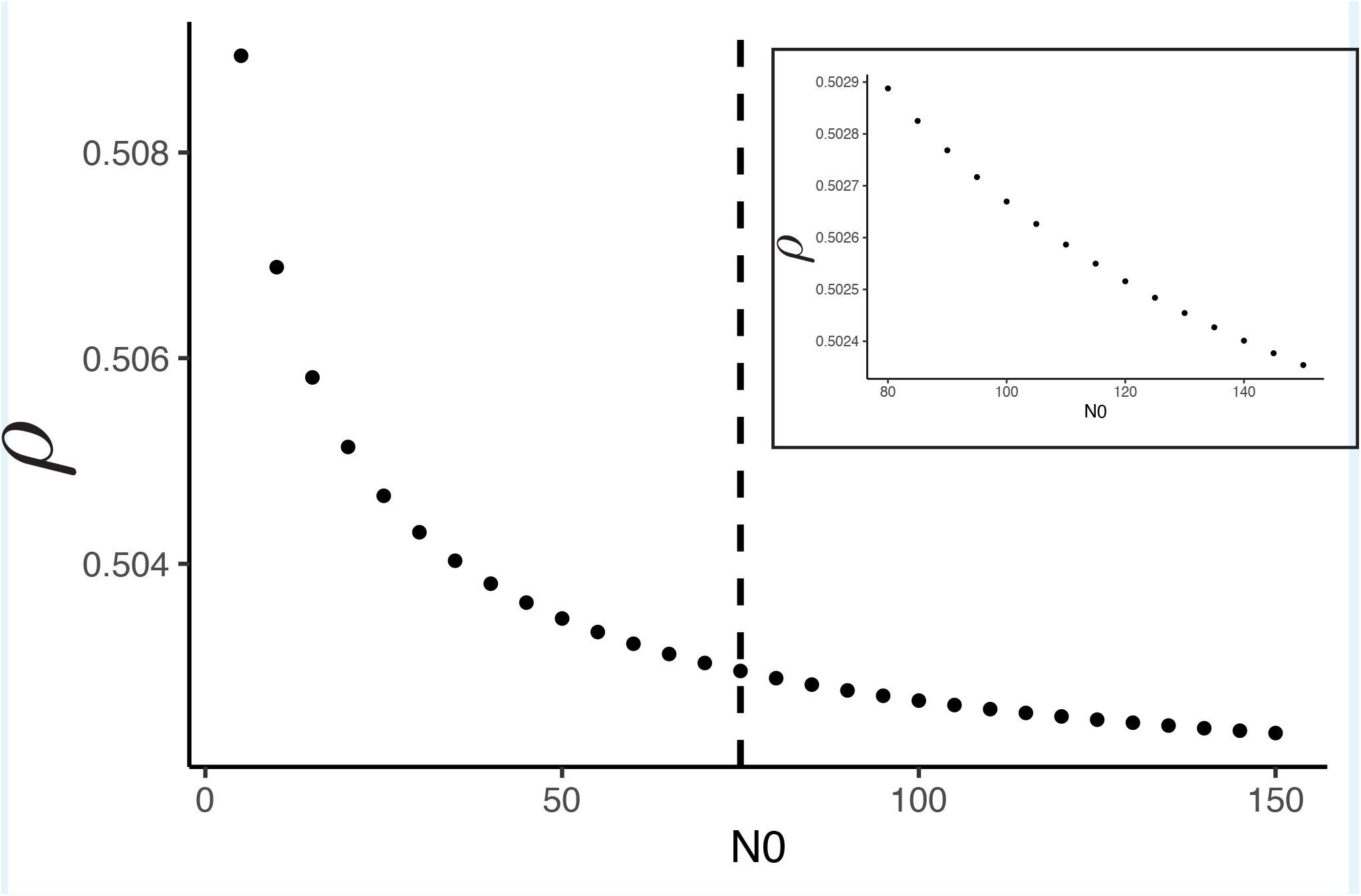
Growth rate bias, *ρ*, for epidemics initialized with different population sizes, *N*_0_, of infected distributed by the equilibrium risk distribution (here *f*_*H*_ = .6. Epidemics were run for 75 populations changes and the vertical line delineates epidemics that stochastic extinction can affect (smaller initial populations). (inset) Focusing on epidemics where not enough population changes have occurred for stochastic extinction to occur.

